# Multi-centre post-implementation evaluation of SARS-CoV-2 antigen-based point of care tests used for asymptomatic screening of continuing care healthcare workers

**DOI:** 10.1101/2021.06.22.21259345

**Authors:** Jamil N Kanji, Dustin T Proctor, William Stokes, Byron M Berenger, James Silvius, Graham Tipples, A Mark Joffe, Allison A Venner

## Abstract

**OBJECTIVES:** Frequent screening of SARS-CoV-2 among asymptomatic populations using antigen-based point of care tests (APOCT) is occurring globally with limited clinical performance data. The positive predictive value (PPV) of two APOCT used in the asymptomatic screening of SARS-CoV-2 among healthcare workers (HCW) at continuing care (CC) sites across Alberta, Canada was evaluated.

**METHODS:** Between February 22 and May 2, 2021, CC sites implemented SARS-CoV-2 voluntary screening of their asymptomatic HCW. Onsite testing with Abbott Panbio or BD Veritor occurred on a weekly or twice weekly basis. Positive APOCT were confirmed with a real-time reverse-transcriptase polymerase chain reaction (rRT-PCR) reference method.

**RESULTS:** A total of 71,847 APOCT (17,689 Veritor and 54,158 Panbio) were performed among 369 CC sites. Eighty-seven (0.12%) APOCT were positive, of which 39 (0.05%) confirmed as true positives using rRT-PCR. Use of the Veritor and Panbio resulted in a 76.6% and 30.0% false positive detection, respectively (p<0.001). This corresponded to a 23.4% and 70.0% PPV for the Veritor and Panbio, respectively.

**CONCLUSIONS:** Frequent screening of SARS-CoV-2 among asymptomatic HCW in CC, using APOCT, resulted in a very low detection rate and a high detection of false positives. Careful assessment between the risks vs benefits of APOCT programs in this population needs to be thoroughly considered before implementation.

## Introduction

Point of Care Testing (POCT), and specifically antigen-based point of care tests (APOCT), represent an important public health measure in managing SARS-CoV-2 transmissions when used appropriately and with acknowledgement of their shortcomings. APOCT is particularly useful in identifying and isolating SARS-CoV-2 positive cases in symptomatic individuals in high-risk settings, such as congregate housing and care facilities or in areas where longer turn-around-times (TAT) of more sensitive nucleic acid testing may compromise containment of transmission. Governments around the world have been quick to support the widescale implementation of APOCT programs [1].

Rapid and wide-scale deployment of APOCT has been promoted despite limited data regarding the analytical and clinical performance of these devices in the various settings in which they have been used. POCT programs that include APOCT have been criticised for the poor sensitivity of these tests compared to PCR tests, particularly in asymptomatic individuals [2]. However, there has been intense interest in testing asymptomatic individuals from a public health perspective, as some estimate that 30 to 40 percent of infectious individuals are asymptomatic. Rapid identification and isolation of these individuals is viewed as highly effective in breaking chains of transmission [1, 3-5]. However, there is concern that APOCT may demonstrate poor specificity when used for the surveillance of asymptomatic individuals, with reports citing up to 60% false positive rates [6, 7].

Routine serial testing of asymptomatic staff in continuing care facilities using APOCT has been implemented as a purportedly important public health measure for early case identification, and prevention of SARS-CoV-2 outbreaks among vulnerable residents [1, 3-5].

While APOCT may represent an important tool in limiting transmission in congregate settings, the negative impacts of a false positive result for staff and the site needs to be considered and further data on the clinical performance of APOCT in continuing care facilities is required.

Our aim was to evaluate the positive predictive value (PPV) of two APOCT (Abbott Panbio™ and BD Veritor™) in a largescale multicenter implementation of asymptomatic SARS-CoV-2 testing of healthcare workers (HCWs) in continuing care facilities.

## Methods

### Setting

Asymptomatic SARS-CoV-2 testing of HCWs in continuing care facilities province-wide was implemented as POCT as part of the provincial pandemic response in the province of Alberta, Canada (population 4.4 million) [8]. A retrospective review of test results was completed for testing completed from February 22, 2021 – May 2, 2021 inclusive. Facilities were assigned (based on availability, oversight, and preference) to use either the Panbio COVID-19 Antigen Rapid Test (Abbott Laboratories, Chicago, USA) or the Veritor COVID-19 Rapid Antigen Test (Becton, Dickson, and Company, Franklin Lakes, USA), herein referred to as APOCT.

Early in the pandemic, the provincial health authority had instituted a continuous masking policy for all continuing care facilities, where all staff were required to continuously wear a medical mask while at work, to be changed should the mask become soiled or wet [9]. Adjunctive eye protection (face shield or goggles) was advised during all episodes of patient care, or continuously if there was a SARS-CoV-2 outbreak declared at the site [10]. HCWs at all facilities were required to complete a ‘fit for work’ symptom questionnaire twice per shift, and immediately leave their shift after informing their manager and arrange COVID-19 testing should they develop symptoms or screen positive [11].

### Asymptomatic testing

Each site received training for a limited number of on-site health care workers to conduct voluntary asymptomatic staff testing of site staff on a weekly basis. Training, operating procedures and medical/operational oversight were provided by Alberta Precision Laboratories (APL) POCT (for sites run by Alberta Health Services [AHS]). Sites external to AHS had vendor-driven training and medical/operational oversight was provided by the site. Staff conducting testing wore personal protective equipment (PPE) consisting of eye protection, surgical mask, gloves, and a splash-resistant gown. Sampling was done using the nasal swab provided in each test kit with testing conducted as per manufacturer’s guidelines [12, 13]. While awaiting results, staff were permitted to continue working. Staff who were symptomatic were not permitted to receive POCT asymptomatic testing and were directed to access COVID-19 testing through public health test centers. At the discretion of individual facilities, staff testing could be increased to twice weekly for non-outbreak related staff at a site where a COVID-19 outbreak had been declared (staff considered linked to the outbreak were not eligible for asymptomatic testing).

Staff receiving positive APOCT results were required to immediately isolate and obtain a confirmatory throat or nasopharyngeal swab test for SARS-CoV-2 with a real-time reverse-transcriptase polymerase chain reaction (rRT-PCR) result at an approved public health test center. rRT-PCR testing was carried out at the provincial Public Health Laboratory (ProvLab) using a validated laboratory-developed assay [14] or a Health Canada approved test at an accredited laboratory. Confirmatory rRT-PCR testing was not conducted on negative SARS-CoV-2 APOCT results. Individuals with invalid APOCT results were referred for rRT-PCR evaluation. APOCT test results were not recorded in the medical chart of HCWs. Any positive APOCT results were considered presumptive positive and were not included in total provincial COVID-19 case counts, using instead the confirmatory rRT-PCR results. All HCWs with a positive APOCT result were able to obtain a follow-up rRT-PCR test.

### Data extraction and analysis

Data recording, organization, and provision of results were provided by Seniors Health and Continuing Care (AHS). Confirmatory rRT-PCR testing results were extracted from the ProvLab laboratory information system. Data were tabulated in Microsoft Excel. Proportional comparisons were done using a two-sample proportions z-test. Continuous variables were compared using Mann-Whitney tests while categorical variables were compared using Chi-square or Fisher’s exact tests. Significance was set at p<0.05. Confidence intervals were calculated using Wilson’s method. Statistical analysis was conducted using StatPlus (AnalystSoft Inc, Alexandria, USA).

## Results

Over the 10-week period, 369/466 (79.18%) provincial continuing care sites participated in the asymptomatic screening program (Table 1). During the study period, 71,847 APOCT were conducted (mean 7,184.7 tests per week; range 615-11648; 17,689 on the Veritor and 54,158 on the Abbott Panbio) (Table 2). A total of 87 (0.12%) APOCT were positive, of which 39 (44.83%) were confirmed to be true positives using rRT-PCR (time duration between APOCT and rRT-PCR confirmation not available). The APOCT positivity rate did not differ significantly across the ten-week period (Table 1; p=0.35).

**Table 1.** Weekly APOCT for asymptomatic staff at continuing care sites across Alberta, Canada.

| Week | Number of reporting sites | Number of tests performed | Positive APOCTs (%)<sup>a</sup> | Invalid APOCTs (%) | Number confirmed by rRT-PCR (%) |
| --- | --- | --- | --- | --- | --- |
| Feb 22-28, 2021 | 11 | 615 | 1 (0.16) | 4 (0.65) | 0 |
| Mar 1-7, 2021 | 115 | 2,446 | 0 | 4 (0.16) | 0 |
| Mar 8-14, 2021 | 188 | 4,374 | 3 (0.07) | 16 (0.37) | 0 |
| Mar 15-21, 2021 | 309 | 8,680 | 8 (0.09) | 29 (0.33) | 1 (12.50) |
| Mar 22-28, 2021 | 293 | 7,603 | 6 (0.08) | 4 (0.05) | 1 (16.67) |
| Mar 29-Apr 4, 2021 | 311 | 8,029 | 9 (0.11) | 13 (0.16) | 5 (55.56) |
| Apr 5-11, 2021 | 317 | 9,599 | 12 (0.13) | 3 (0.03) | 8 (66.67) |
| Apr 12-18, 2021 | 325 | 9,312 | 15 (0.16) | 4 (0.04) | 6 (40.00) |
| Apr 19-25, 2021 | 321 | 9,541 | 17 (0.18) | 6 (0.06) | 10 (58.82) |
| Apr 26-May 2, 2021 | 369 | 11,648 | 16 (0.14) | 4 (0.03) | 8 (50.00) |
| Total | 369 | 71,847 | 87 (0.12) | 87 (0.12) | 39 (44.83) |

Abbreviations: Apr – April; Feb – February; Mar – March; rRT-PCR – real time reverse transcriptase polymerase chain reaction.
<sup>a</sup>Change in APOCT positivity across the ten weeks is not significant (Pearson's Chi-square, p=0.35).

**Table 2.** APOCT and rRT-PCR confirmatory results for asymptomatic continuing care health care workers in Alberta, Canada.

| Item | BD Veritor | Panbio | Total (%) |
| --- | --- | --- | --- |
| Total APOCTs | 17,689 | 54,158 | 71,847 |
| Positive APOCTs (%) | 47 (0.27) | 40 (0.07) | 87 (0.12) |
| Negative APOCTs (%) | n/a | n/a | 71,673 (99.76) |
| Invalid APOCTs (%) | n/a | n/a | 87 (0.12) |
| Positive APOCTs confirmed by rRT-PCR (%) | 11 (0.06) | 28 (0.05) | 39 (0.05) |
| Number of false positives (%) | 36/47 = 76.60%<sup>a</sup> | 12/40 = 30.00%<sup>a</sup> | 48/87 = 55.17% |
| Positive predictive value (%) | 11/47 = 23.40%<sup>a</sup> | 28/40 = 70.00%<sup>a</sup> | 39/87 = 44.83% |

Abbreviations: n/a – not available
<sup>a</sup>Comparison between BD Veritor and Abbott Panbio: $p < 0.0001$ .

Compared to the Panbio, the false detection rate was significantly higher using the Veritor (76.6%, 95% confidence interval (CI) 62.8-86.4% vs 30.0%, 95%CI 18.1-45.4%; p<0.0001) (Table 2). The PPV was significantly higher with the Panbio test (70%, 95%CI 54.6-81.9% vs 23.4%, 95%CI 13.6-37.2%; p<0.0001).

During this time, a total of 66,338 cases of COVID-19 were diagnosed in Alberta (prevalence of 1.5%) with an average rRT-PCR positivity rate (for all specimens tested) of 7.2% (median 6.3%; range 3.8-13.6%) [15]. During the study period, a total of 32 outbreaks were declared in continuing care facilities across the province (one in March 2021, 29 in April 2021, and two in May 2021). HCWs in continuing care facilities were offered mRNA COVID-19 vaccines starting December 16, 2020 alongside the residents in these facilities (dosing between first doses being four and six weeks for residents and HCWs respectively). The proportion of all HCWs working in Alberta continuing care facilities who had received one dose of COVID-19 vaccine as of February 1 and May 2, 2021 was 1.1 and 69.9% respectively. The proportion having received two doses of vaccine by these dates was 16.0 and 83.6% respectively. The demographics and proportions of vaccinated HCWs tested were not available, as neither were the follow-up rRT-PCR testing results of 87 individuals with invalid APOCT results.

## Discussion

Despite great emphasis on the use of APOCT as a tool to combat the COVID-19 pandemic, expansion of APOCT for asymptomatic testing has been met with hesitation due to their limited PPV, especially in settings of low disease prevalence [16]. This study confirms and quantitates these concerns, demonstrating in a large-scale implementation of asymptomatic screening in continuing care HCWs, that APOCT have a low overall PPV (44.83%) and a high proportion of false positives (55.17%).

Similar performance concerns related to APOCT have been raised by a number of studies. A recent Cochrane Database systematic review reported the overall sensitivity (Sn) and specificity (Sp) of APOCT in symptomatic disease to be 72.0 and 99.5% (twenty-seven studies combined) and asymptomatic disease to be 58.1% and 98.9% respectively (twelve studies combined) [2]. Applying this to the provincial prevalence of disease we observed during the study period (1.5%), indicates our findings are consistent, with an expected PPV for APOCT around 44.6%. Several studies have reported a PPV as low as 33.3% [17, 18].

Many reports of asymptomatic SARS-CoV-2 testing campaigns using either APOCT or rRT-PCR tests have demonstrated low test positivity rates, suggesting high volume testing of asymptomatic individuals is of low yield [19-22]. Another Canadian study evaluating universal APOCT screening of HCWs in continuing care also found a similar APOCT positivity rate of 0.16% [23]. The prevalence of positive results in our cohort was similar at 0.12%, however the true prevalence of disease in our cohort is much less if true positives are accounted for (0.05%). Therefore, when considering the true positivity rate, the yield of asymptomatic screening programs is poor.

The Veritor displayed both a lower PPV and a higher proportion of false positive tests compared to the Panbio. Both tests were initially authorised by the FDA and Health Canada for the diagnosis of COVID-19 within the first five (Veritor) to seven (Panbio) days of COVID-19 symptom onset [16]. Our findings are consistent with the literature, demonstrating the PPVs for the Veritor in asymptomatic individuals ranging from 24.9-50.1% (with disease prevalence of 0.5-1.5%) [24] and that of the Panbio to be 28.0-54.0%, using prevalence estimates of 0.5-1.5% [24-26]. Thus, the Panbio would be preferred over the Veritor for asymptomatic screening of COVID-19.

The impact of a false positive result can lead to considerable burden and distress on both individuals and the health care system. False positive test results in HCWs and others can lead to loss of income due to the need to isolate, collateral effects to close contacts, and also psychological damage due to misdiagnosis, stigma and fear of infecting others (especially loved ones who could have poor outcomes) [27]. In health care settings, the need for isolation in response to a false positive result can lead to staff shortages, adding further stress on other employees and patients. False positive APOCT results in acute care settings have resulted in unnecessary cancellation or postponement of treatment/procedures, and also risks that individual possibly being exposed to SARS-CoV-2 if moved into a COVID-19 treatment unit based on the APOCT result [28]. Thus, we strongly recommend proper education about the meaning of APOCT results, and ensuring processes are in place for timely confirmation with rRT-PCR if APOCT is implemented.

The setup of asymptomatic testing programs using APOCT requires considerable input of resources and logistical organization. To ensure the utmost quality of testing, APOCT should undergo appropriate test verification, creation of standard operating procedures and training programmes, and include appropriate medical and operational oversight through an accredited laboratory when implementing the associated POCT program [29]. Information technology specialists are generally required to establish reporting systems so that results can be accurately captured to facilitate public health reporting, disease notification, and contact tracing. Furthermore, ongoing oversight to address reporting errors, test distribution, and ongoing test validation must be built into the APOCT infrastructure [30]. The impact of false positive results, alongside these important operational considerations must be accounted for when deciding on the benefit of an APOCT screening program.

The principal limitation of this study is that only positive APOCT tests were confirmed by rRT-PCR, and thus no other test parameters other than a PPV could be calculated. The data however, still confirms in a large asymptomatic HCW population, that APOCT has a low PPV. The major strengths of this study lie in the large number of tests performed, participating sites, and inclusion of two different APOCT platforms.

The utility of screening asymptomatic individuals with COVID-19 APOCT is best determined by weighing the costs and benefits specific to each setting. With widespread vaccination increasing and disease prevalence falling, fewer positive test results are likely to be found among asymptomatic persons in the coming months (with a large proportion of them being false-positives), leading such endeavors to be quite costly from a materials and human resources perspectives for most health care systems. Therefore, it is important to reconsider the value and effectiveness of asymptomatic APOCT COVID-19 programs over the long term, especially in the context of increasing population immunity.

## Data Availability

Due to confidentiality issues, data is available upon reasonable request.

## Declarations

### Ethics approval

Presentation of the data contained in this report has been approved by the Human Research Ethics Board at the University of Alberta (study identifier Pro00110831).

## Acknowledgements

We would like to thank the Alberta Precision Laboratories POCT staff for their support to the Panbio POCT program who developed and supported designated continuing care facilities. We would also like to thank the staff within the Provincial Seniors Health program who developed the program parameters and operationalised the program (Sandra Gugins and Sharon Leontowicz), and participating Continuing Care sites, staff responsible for the asymptomatic testing programs and those staff who participated in screening.

## CRediT Author Statement

Jamil N Kanji – formal analysis, data curation, writing – original draft, writing – review and editing, visualization

Dustin Proctor – validation, writing – original draft, writing – review and editing

William Stokes – conceptualization, methodology, validation, investigation, resources, project administration, writing – original draft, writing – review and editing

Byron Berenger – conceptualization, validation, resources, writing – original draft, writing – review and editing

James Silvius - conceptualization, validation, resources, project administration, writing – review and editing

Graham Tipples - conceptualization, validation, resources, project administration, writing – review and editing

Mark A Joffe - conceptualization, validation, resources, project administration, writing – review and editing

Allison Venner – conceptualization, methodology, validation, investigation, resources, project administration, writing – original draft, writing – review and editing, supervision

### Availability of data and materials

The datasets used and/or analysed during the current study are available from the corresponding author on reasonable request.

## STROBE CHECKLIST

**Source of STROBE checklist: https://www.strobe-statement.org/fileadmin/Strobe/uploads/checklists/STROBE_checklist_v4_cohort.pdf**

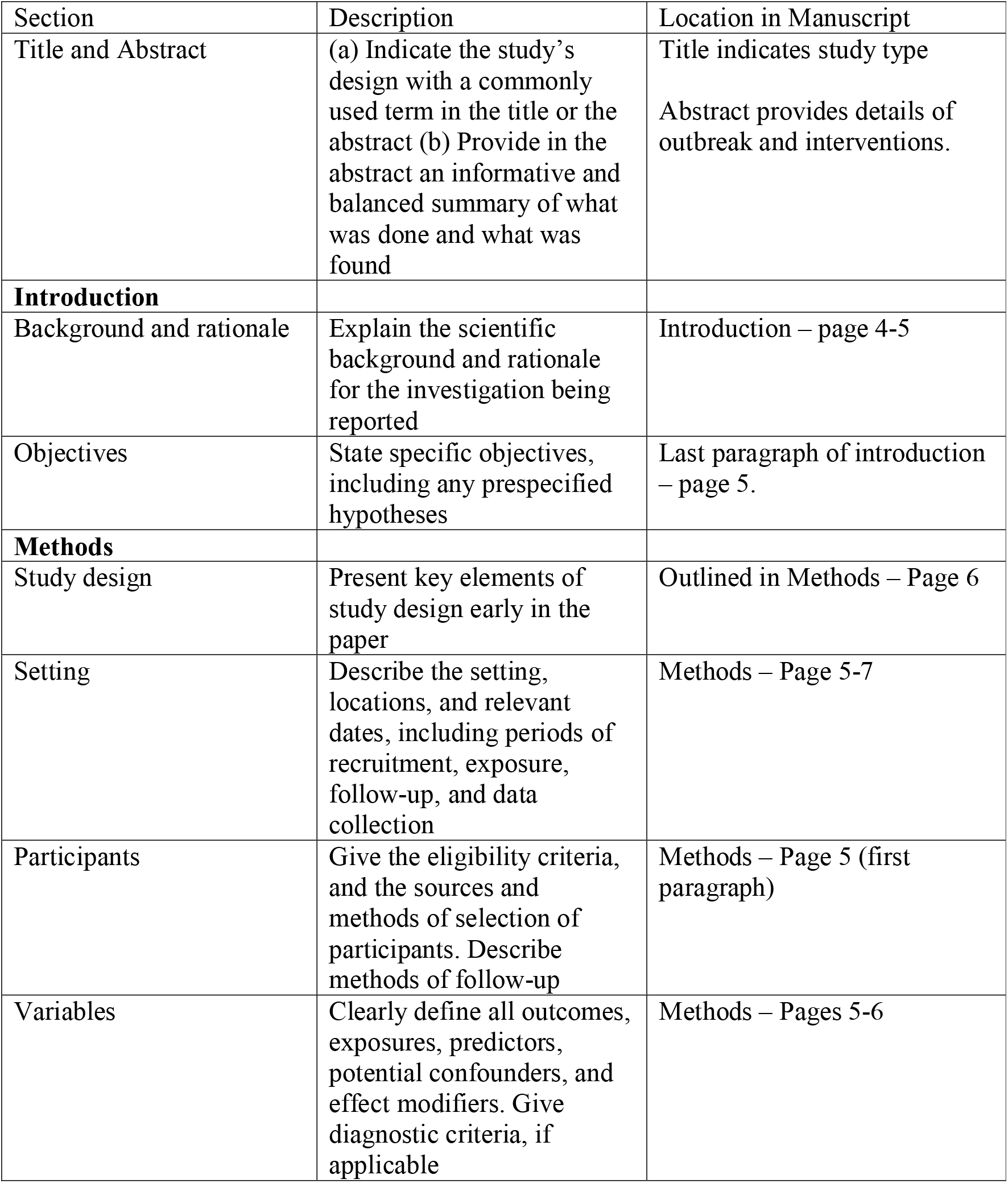
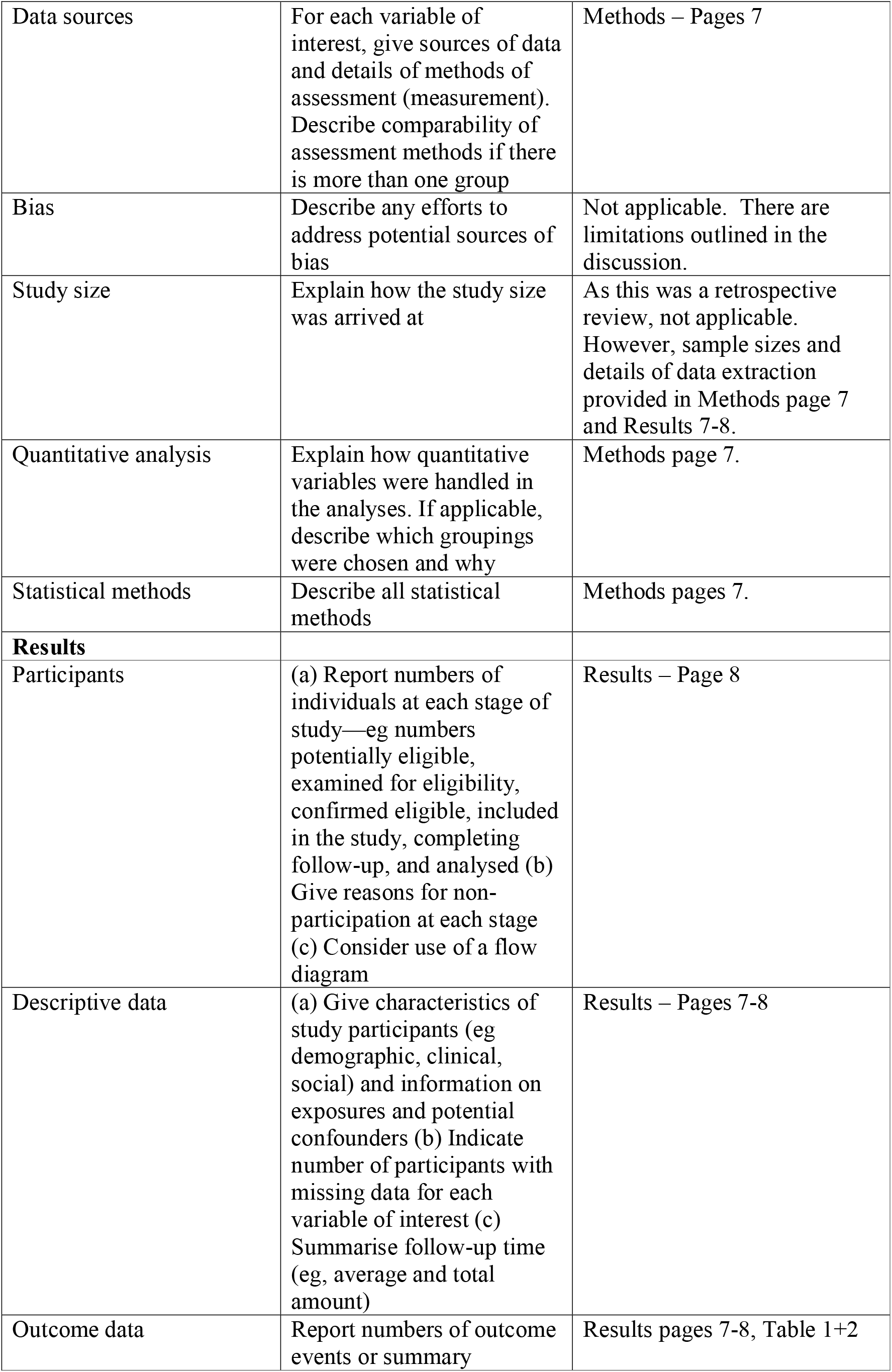
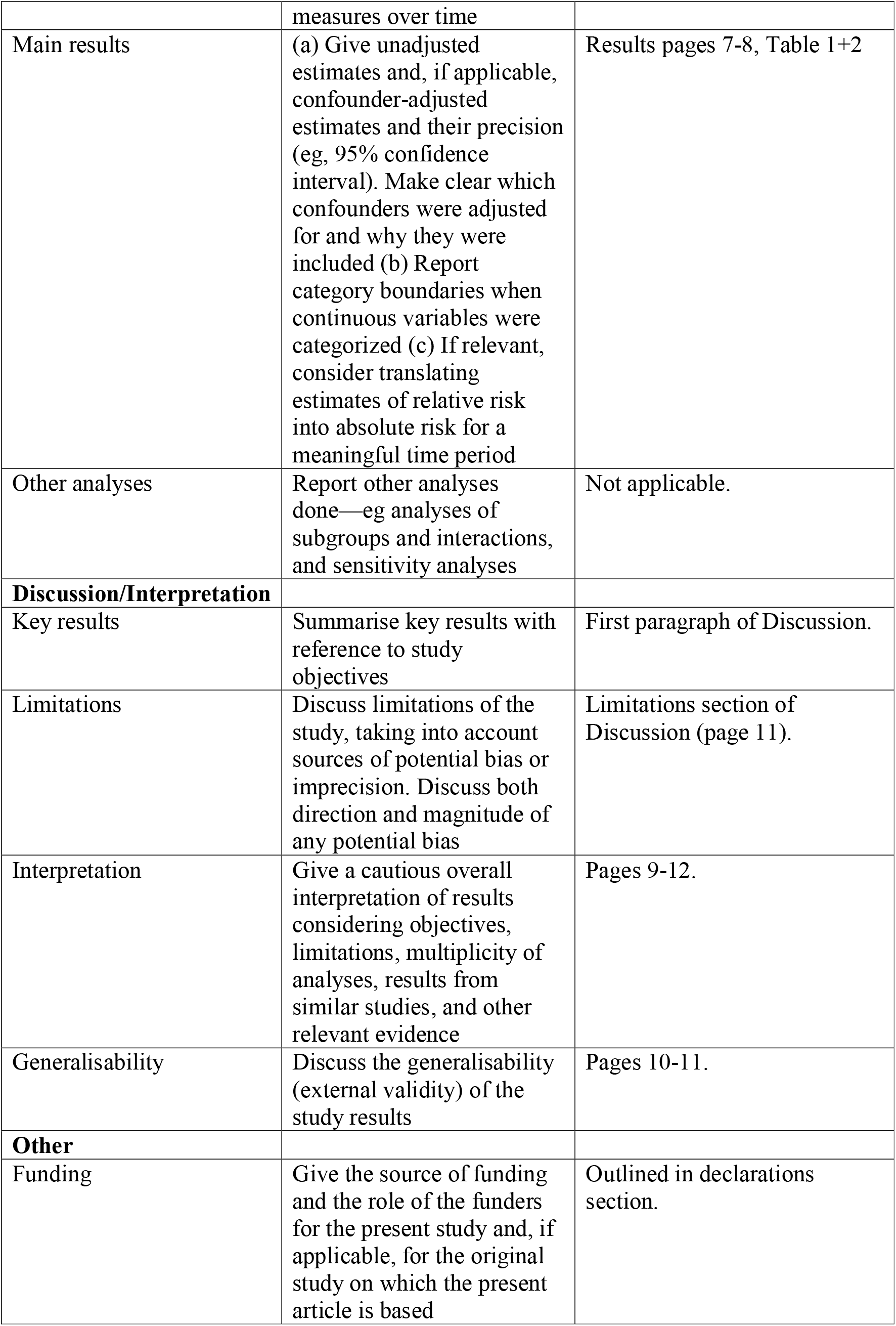

